# Analysis of dietary vitamin C intake levels and the risk of hyperuricemia and gout based on cross-sectional studies and bi-directional Mendelian randomisation

**DOI:** 10.1101/2024.01.18.24301481

**Authors:** Zi-Ning Peng, Xing-Qiang Wang, Qian Deng, Wei-Tian Yan, Wei-Qing Zhao, Yong-Bin Pu, Nian Liu, Ling-Li Gu, Jiang-Yun Peng

## Abstract

**Background:** Vitamin C, a common antioxidant, may be useful for treating hyperuricemia and gout. The aim of this study was to investigate the risk association between dietary vitamin C intake and hyperuricemia/gout and to test for causality using the bi-directional Mendelian randomisation method.

**Methods:** Cross-sectional studies were selected from the National Health and Nutrition Examination Survey from 2007–2018 to assess the association between dietary vitamin C intake and the risk of hyperuricemia/gout, according to multivariate logistic regression modelling. Bi-directional Mendelian randomisation studies were conducted using genetic data from large-scale genome-wide association surveys of supplemental vitamin C, pharmacological vitamin C, and ascorbic acid intake and hyperuricemia/gout; these aimed to infer causal relationships between vitamin C and hyperuricemia/gout. Inverse variance weighting was used as the primary method of Mendelian randomisation analysis. A series of sensitivity analyses were used to assess multiplicity.

**Results:** The cross-sectional study included 17.52% and 82.48% patients with and without hyperuricemia, respectively, as well as 2.67% and 97.33% patients with and without gout, respectively. In the model correcting for all covariates, the association between vitamin C intake and the risk of hyperuricemia/gout was stable, and the risk of hyperuricemia was generally lower in patients who consumed >111.75 mg vitamin C than in other patients when comparing three models with different moderators. Restricted cubic spline scores indicated that vitamin C intake recommendations of 75–525 mg and 75–225 mg were effective for targeting hyperuricemia and gout, respectively. In inverse variance weighting in the Mendelian randomisation analysis, the amount of vitamin C absorbed was negatively associated with hyperuricemia (OR = 0.985, 95% CI = 0.973–0.997, p = 0.015), and supplemental vitamin C was negatively associated with gout (OR = 0.857, 95% CI = 0.797–0.921, p < 0.001); sensitivity analysis yielded consistent results. Reverse Mendelian randomisation analysis showed that vitamin C had no reverse causal relationship with hyperuricemia and gout.

**Conclusion:** We hypothesise that a dietary vitamin C intake of 75–525 mg or 75–225 mg may reduce the risk of hyperuricemia and gout, respectively. Further research with larger samples is required to confirm this.

## 1 Introduction

Gout has long been known as the “disease of kings” because of the disproportion rates of gout in royal families (1). In modern society, hyperuricemia (HUA) and other musculoskeletal diseases are common medical issues due to improvements in living standards (2). Cases of HUA and gout are increasing globally, particularly in higher-income, developed economies. Although current epidemiological data are incomplete, approximately one in five people are affected by increased uric acid levels and their effects; the prevalence of HUA and gout are approximately 20% and 2.6%, respectively (3, 4).

Generally, gout begins as asymptomatic HUA, gradually developing into clinical symptoms accompanied by uric acid crystal deposits in the joint cavities, and finally advancing to a diagnosis of gout. This continuous process produces a series of complications (5). Persistent HUA can easily trigger end-stage diseases, resulting in increased mortality. Therefore, controlling high uric acid levels is crucial in determining treatment opportunities for gout. Epidemiological research shows that behavioural factors, especially dietary factors, affect uric acid metabolism. Although several anti-gout medications, such as febuxostat and colchicine, have been approved for use by the US Food and Drug Administration in recent years, these medications have significant adverse effects, including renal and hepatic failure and cardiotoxicity (6). Therefore, vitamin C (VC) has piqued the interest of researchers as a common antioxidant.

VC, also known as ascorbic acid, plays a pleiotropic role in living organisms. In 1932, Hungarian biologist Albert Szent-Györgyi first discovered VC and used it to regulate different biological processes (7). VC possesses bi-directional properties, acting as both an antioxidant and a pro-oxidant. Additionally, it exhibits properties that are related to the immune system. These characteristics of VC can aid in reducing HUA (8). Although dietary approaches appear to play an important role in preventing gout attacks or the onset of the disease, a comprehensive understanding of the interaction between uric acid and VC in the human body remains insufficient.

In HUA- or gout-related research, it is already very common to observe the protective properties of VC. For example, research shows that dietary and supplemental VC intake are negatively correlated with HUA risk (9). However, focusing solely on HUA is insufficient; therefore, our research also included participants with gout. In addition, the National Health and Nutrition Examination Survey (NHANES) 2007-2018 was used to add more data compared to previous research, and quartiles of dietary VC were observed. Supplemental VC was not considered as part of the overall dietary intake to avoid potential disruptions in our analysis; therefore, the effects of supplemental VC were excluded from our study.

Mendelian randomisation (MR) is an epidemiological method that uses genetic variation as an instrumental variable (IV) to represent exposure to a variable of interest. It is also used to investigate the effect of exposure on specific outcomes (10). In the present study, we investigate the underlying causal relationship between VC and gout/HUA. We performed MR analysis using genome-wide association study (GWAS) data. MR is the ideal tool for studying immune diseases because when exposed to external environmental factors, genetic variables can affect life expectancy. We used the bi-directional MR method to analyse and predict the causal effects of VC on gout and HUA, which helped mitigate bias from dietary questionnaires in the NHANES database, thereby enhancing the reliability of the results.

## 2 Material and Methods

This study consisted of two parts. In the first part, we conducted a study using data from a large sample of the NHANES to explore the association between VC intake and HUA/gout. Due to questionnaire bias in the NHANES database, we supplemented the second part with a bi-directional MR analysis utilising GWAS pooled statistics to reduce bias and avoid reverse causality and assessed the causal effects of VC intake on HUA and gout. The overall study design is shown in Figure 1.

**Figure 1.**
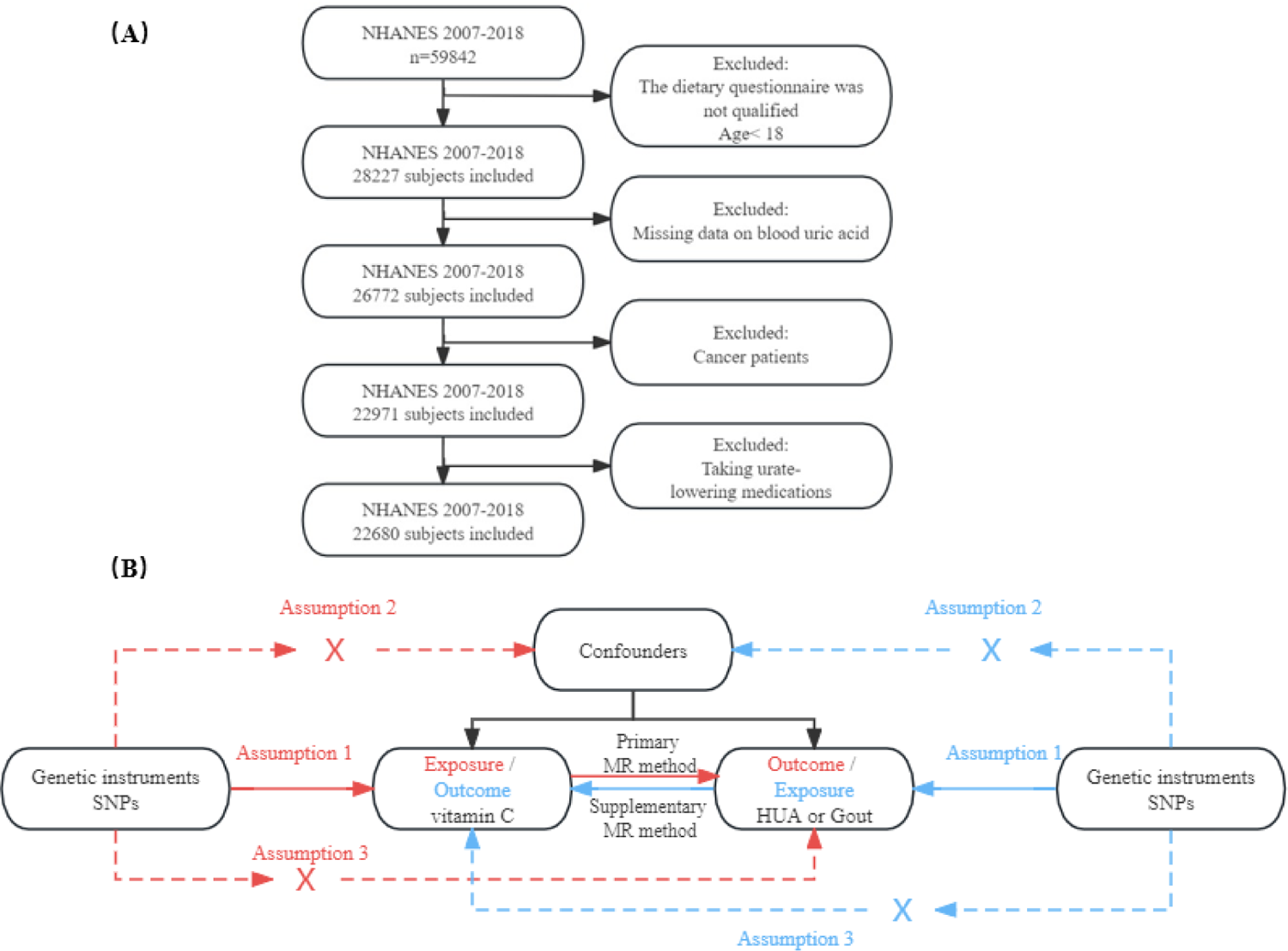
Study design overview, Note. (A) Flow chart of NHANES 2007-2018 sample selection, (B) General design of the MR Study.

### 2.1 Cross-sectional Study

#### 2.1.1 Study Population

NHANES examined a nationally representative sample of the civilian non-institutionalised population of the USA of all ages using a complex multistage probability design. We used the most recent NHANES database containing data from six cycles (2007–2018) (n = 59,842). Figure 1A shows the NHANES 2007–2018 sample selection flowchart. Following a home interview, participants were invited to complete additional questionnaires, undergo testing, and provide blood and other biospecimens in a mobile examination centre. The National Centre for Health Statistics Research Ethics Review Board approved NHANES, and all participants provided informed consent for the home interview and tests at the mobile examination centre. There were 28,227 participants aged 18 years or older who were eligible for the dietary questionnaire in this survey. Individuals who did not provide a blood uric acid specimen (n = 1,455), those who currently or previously had a tumour (self-reported) (n = 3,801), and those taking uric acid-lowering medications (allopurinol, febuxostat, or probenecid) that interfered with the investigation (n = 291) were excluded, and the remaining 22,680 individuals were enrolled in our next step of analysis.

#### 2.1.2 Dietary VC Intake

Dietary VC intake was assessed using 24 h recall. Relying on the US Department of Agriculture Food and Nutritional Database (2007–2018) to determine the nutrient composition, supplied energy, and other components of diet and fluids, all participants underwent two 24 h recalls. The interviews included all types of food and fluids consumed. Thereafter, we analysed the average VC intake, including VC from all meals consumed by the participants, while excluding VC from supplements.

#### 2.1.3 HUA and Gout Diagnosis

Serum concentrations of uric acid were measured using the Beckman UniCel® DxC 800 Synchron or Beckman Synchron LX20 (Beckman Coulter, Inc., Brea, CA, United States) after the uric acid was oxidised by uricase to form allantoin and H_2_O_2_. HUA was diagnosed when uric acid levels were ≥420 μmol/l (7 mg/ dL) in men and ≥360 μmol/l (6 mg/dL) in women (11). Gout flares were defined by self-assessment in a questionnaire.

#### 2.1.4 Covariates

Demographic characteristics included age, sex, race (Hispanic American, non-Hispanic white, non-Hispanic black, and other races), marital status (married, widowed, divorced, and unmarried), and education level (less than high school, high school graduate, and equal to college and above). Other covariates included smoking status (never, ex-smoker, and current persistent smoker), alcohol use (defined as at least one drink per month in the last year), poverty level, body mass index, and waist circumference. History of cardiovascular disease, hypertension, hyperlipidaemia, kidney stones, and diabetes mellitus were defined as self-reported medical diagnoses. Glomerular filtration rate was used to assess renal function, which was calculated using the CKD-EPI creatinine equation (12). Blood specimens were processed, stored, and shipped to their respective laboratories for analysis, including creatinine, glucose, cholesterol, triglyceride, direct cholesterol, and uric acid analysis.

#### 2.1.5 Statistical Analysis

R version 4.2.2 was used for all analyses. Counts and proportions were used for categorical variables, whereas means and standard deviations were used for continuous data, and sampling weights were considered for all analyses. For two-tailed tests, p < 0.05 was considered statistically significant. Based on VC intake quartiles, subjects were categorised into Q1 (0–29.1 mg), Q2 (29.1– 61.0 mg), Q3 (61.0–111.75 mg), and Q4 (>111.75 mg), and descriptive statistics were used to analyse the characteristics of the participants. Categorical variables were compared using the Chi-square test, and continuous variables were compared using ANOVAs or the Kruskal–Wallis H test. In this study, three models were produced: Model 1 included all variables, Model 2 corrected for some demographically relevant variables, and Model 3 corrected for all covariates. Logistic regression models and restricted cubic spline analyses were carried out to assess the correlation between dietary VC intake with the risk of developing gout and HUA.

### 2.2 Bi-directional MR Study

#### 2.2.1 Study Design

The overall design of the MR study is shown in Figure 1B. To obtain valid causal effect estimates, IVs in the forward MR model need to meet the following three conditional assumptions: 1) genetic variations are significantly associated with VC intake levels; 2) genetic variations are not associated with other confounders; and 3) they affect gout or HUA only through the VC intake levels. Inverse MR model IVs need to meet the following three conditional assumptions: 1) genetic variations are significantly associated with gout or HUA; 2) genetic variations are not associated with other confounders; and 3) they affect VC intake levels only through gout or HUA.

#### 2.2.2 Data Sources and Genetic Tools

Genetic tools were mainly derived from the latest GWAS meta-analysis, i.e. GWAS summary data (13). GWAS is a genome-wide test of polymorphic genetic variants (markers) in multiple individuals, enabling researchers to obtain genotypes for statistical analysis at the population level. We used GWAS to identify sequence variants, known as single nucleotide polymorphisms (SNPs), present on a genome-wide scale in humans. From this, we screened for SNPs associated with HUA, gout, and VC intake (14). HUA and gout data were obtained from the EBI database in the GWAS database, and VC data were obtained from the UKB database. Due to the lack of GWAS data on dietary VC, for the sake of completeness, we used all VC-related data from the UKB database for MR analysis, including data on “supplements: vitamin C”, “medication: vitamin C” and “vitamin C absorbated”. The specific data examined in this study are shown in Supplementary Table 1.

#### 2.2.3 Statistical Analysis

MR analysis was mainly performed using inverse variance weighting (IVW)-random effects and IVW-fixed effects models. All statistical analyses were performed using “two sample MR” in R software. In addition, weighted median and MR-Egger regression were applied to supplement the analyses. Heterogeneity and pleiotropy among the selected SNPs were assessed using Cochrane Q-analysis and MR-Egger intercepts. Leave-one-out analysis was performed to test whether the estimates were driven by individual SNPs. Statistical power and F-statistic computational measurements ensured sufficient statistical efficacy to avoid weak IV bias. Power calculation was performed using the online tool mRnd^1^ and the IV was deleted when the IV strength measure F-statistic was <10 to minimise the chance of MR analysis bias. Finally, reverse MR was applied to test for the presence of reverse causal links.

## 3 Results

### 3.1 Cross-sectional Study

#### 3.1.1 Basic Characteristics of Study Participants

Tables 1 and 2 summarise the overall characteristics of the study participants, that is, whether they had HUA and gout, respectively. Among the participants, 17.52% had HUA and 82.48% did not have HUA, whereas 2.67% had gout and 97.33% did not have gout. Separate analyses were performed according to sex, race, marital status, education, alcohol consumption, smoking status, hypertension, hyperlipidaemia, and diabetes, in relation to the presence of HUA or gout. Table 1 shows that, with the exception of alcohol consumption (p = 0.8) and poverty level (p = 0.2), all other variables exhibit statistically significant differences (p < 0.001) between individuals with HUA and those with kidney stones (p = 0.006). Table 2 shows that, in addition to education level (p = 0.4), poverty level (p = 0.4), and cholesterol (p = 0.6), race (p = 0.014), alcohol consumption (p = 0.006), and VC (p = 0.003) showed statistical significance with regard to having gout. Furthermore, the other covariates showed statistically significant differences with regard to having gout (p < 0.001). The overall basic characteristics indicated that the prevalence of HUA and gout was higher in men than in women, and that patients with HUA and gout had lower VC intake than patients without these diseases.

**Table 1.**
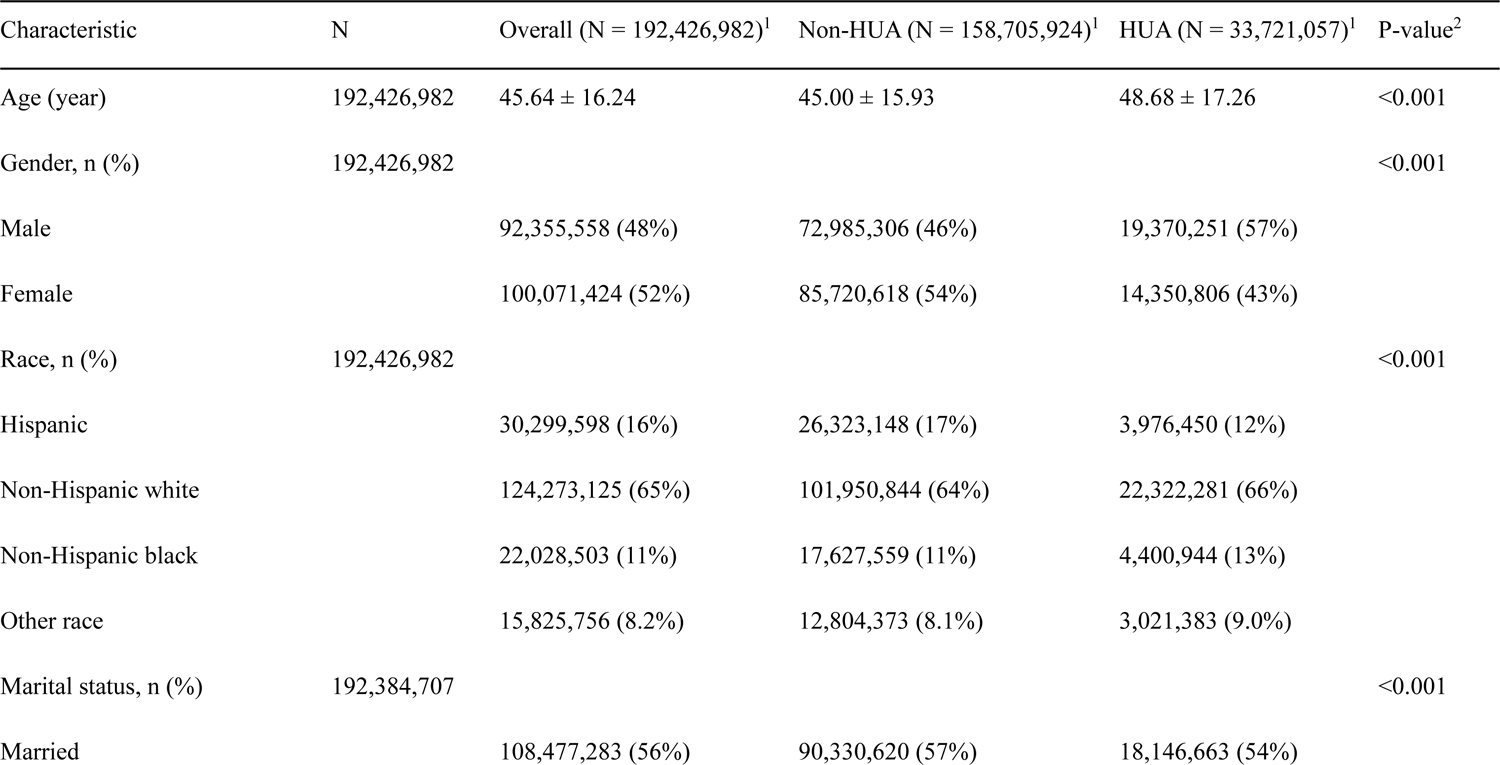
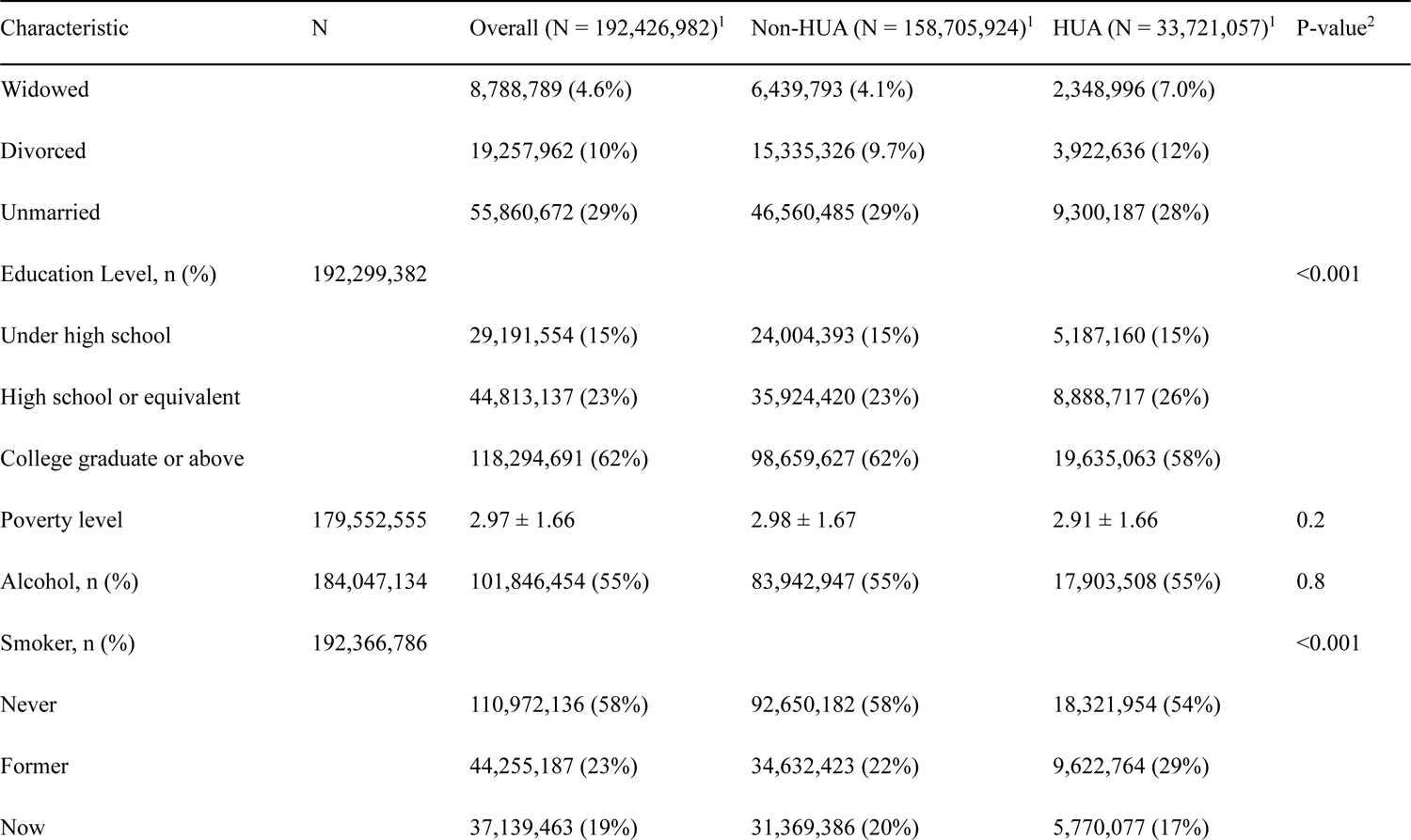
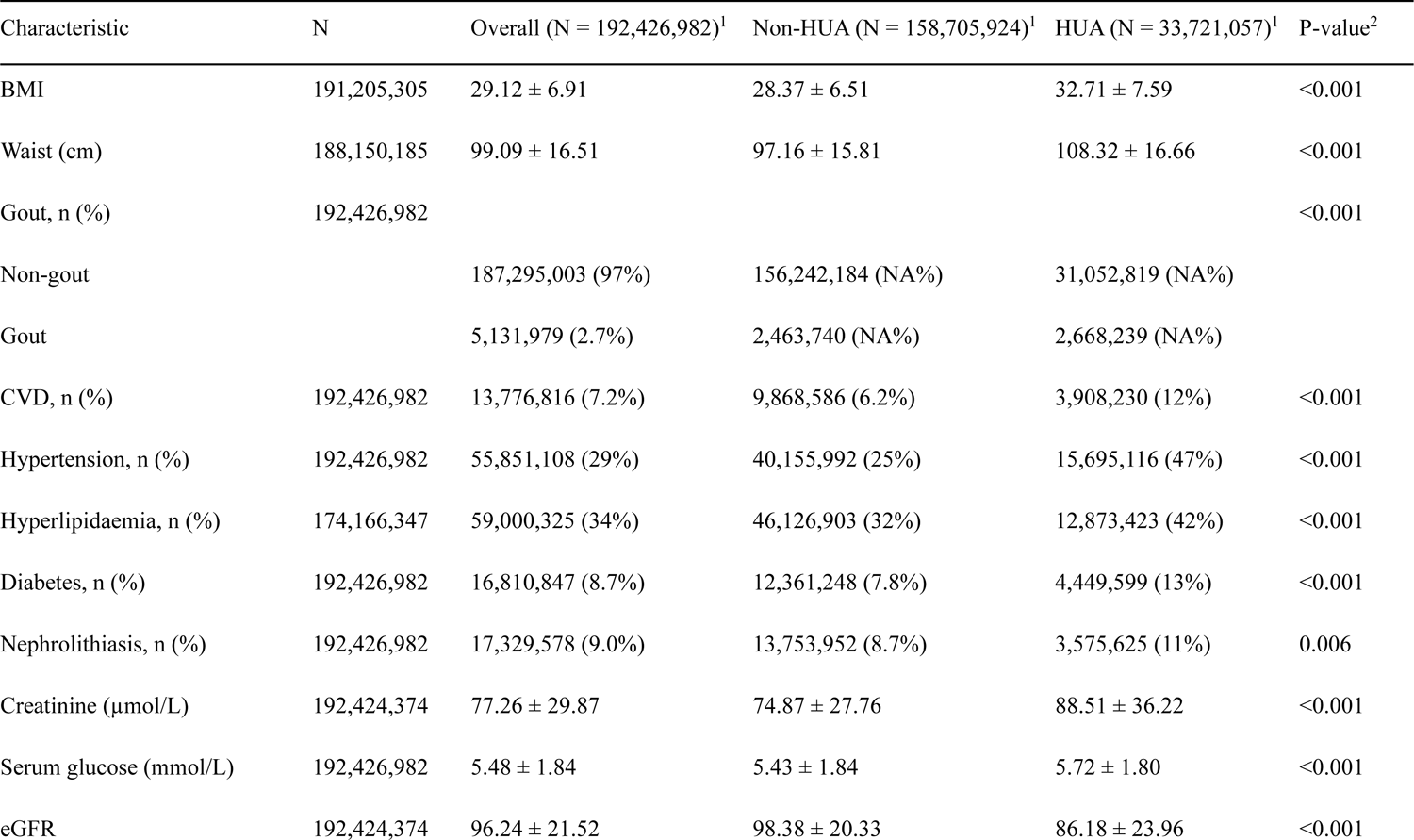
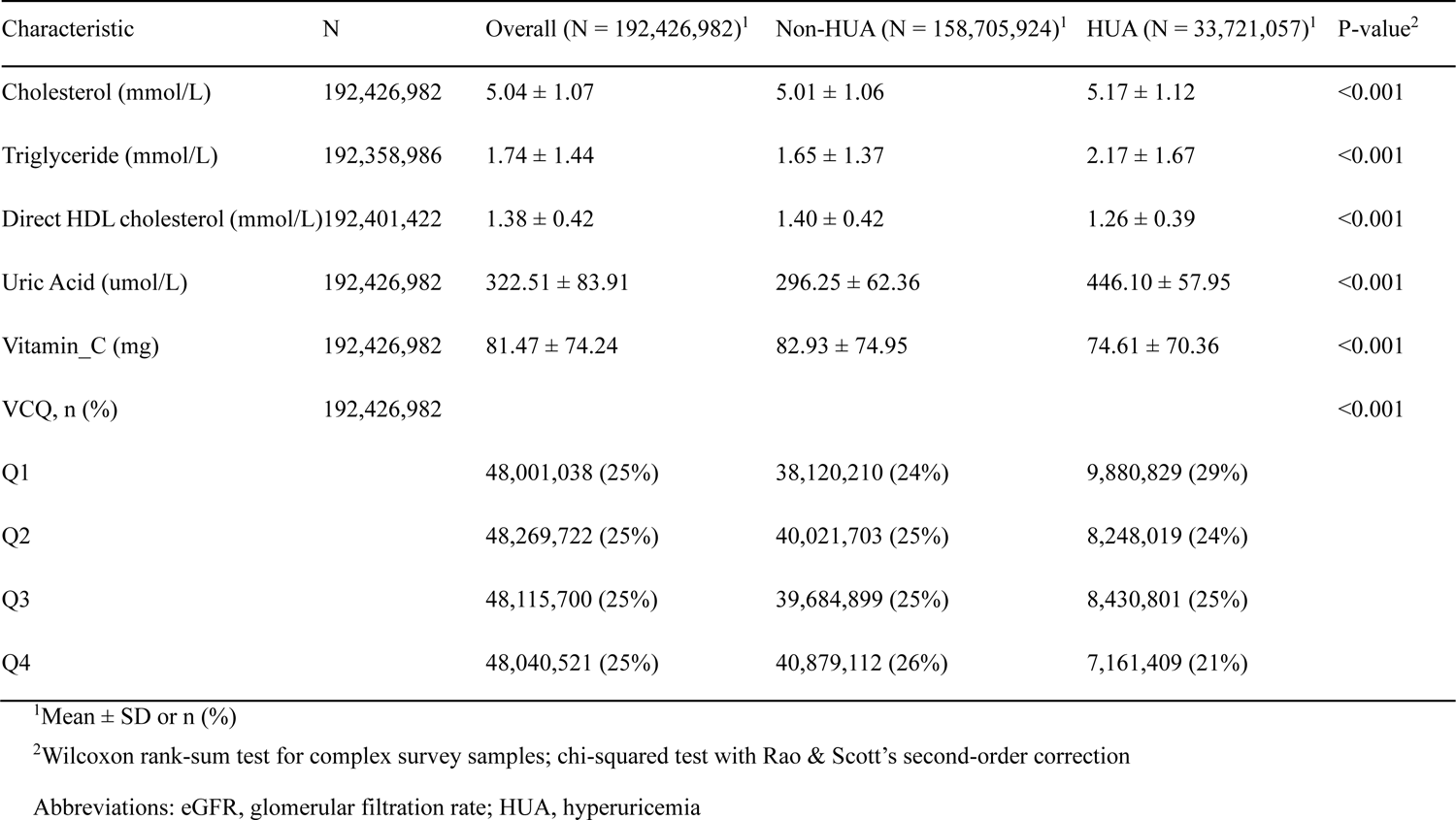
Basic characteristics of participants in the HUA study.

**Table 2.** Basic characteristics of participants in the gout study.

| Characteristic | N | Overall (N = 192,426,982)<sup>1</sup> | Non-gout (N = 187,295,003)<sup>1</sup> | Gout (N = 5,131,979)<sup>1</sup> | P-value<sup>2</sup> |
| --- | --- | --- | --- | --- | --- |
| Age (year) | 192,426,982 | 45.64 ± 16.24 | 45.32 ± 16.19 | 57.55 ± 12.94 | <0.001 |
| Gender, n (%) | 192,426,982 |  |  |  | <0.001 |
| Male |  | 92,355,558 (48.0%) | 88,878,925 (47.5%) | 3,476,633 (67.7%) |  |
| Female |  | 100,071,424 (52.0%) | 98,416,078 (52.5%) | 1,655,346 (32.3%) |  |
| Race, n (%) | 192,426,982 |  |  |  | 0.014 |
| Hispanic |  | 30,299,598 (15.7%) | 29,834,570 (15.9%) | 465,027 (9.1%) |  |
| Non-Hispanic white |  | 124,273,125 (64.6%) | 120,723,187 (64.5%) | 3,549,938 (69.2%) |  |
| Non-Hispanic black |  | 22,028,503 (11.4%) | 21,380,418 (11.4%) | 648,085 (12.6%) |  |
| Other race |  | 15,825,756 (8.2%) | 15,356,828 (8.2%) | 468,928 (9.1%) |  |
| Marital status | 192,384,707 |  |  |  | <0.001 |
| Married |  | 108,477,283 (56.4%) | 105,141,701 (56.1%) | 3,335,582 (65.0%) |  |
| Widowed |  | 8,788,789 (4.6%) | 8,457,078 (4.5%) | 331,711 (6.5%) |  |
| Divorced |  | 19,257,962 (10.0%) | 18,523,213 (9.9%) | 734,749 (14.3%) |  |
| Unmarried |  | 55,860,672 (29.0%) | 55,133,311 (29.4%) | 727,361 (14.2%) |  |
| Education Level, n (%) | 192,299,382 |  |  |  | 0.4 |
| Under high school |  | 29,191,554 (15.2%) | 28,298,957 (15.1%) | 892,597 (17.4%) |  |
| High school or equivalent |  | 44,813,137 (23.3%) | 43,579,805 (23.3%) | 1,233,333 (24.0%) |  |
| College graduate or above |  | 118,294,691 (61.5%) | 115,288,641 (61.6%) | 3,006,049 (58.6%) |  |
| Poverty level | 179,552,555 | 2.97 ± 1.66 | 2.97 ± 1.66 | 2.88 ± 1.68 | 0.4 |
| Alcohol, n (%) | 184,047,134 | 101,846,454 (55.3%) | 99,515,325 (55.6%) | 2,331,129 (46.8%) | 0.006 |
| Smoker, n (%) | 192,366,786 |  |  |  | <0.001 |
| Never |  | 110,972,136 (57.7%) | 108,756,367 (58.1%) | 2,215,769 (43.2%) |  |
| Former |  | 44,255,187 (23.0%) | 42,344,158 (22.6%) | 1,911,029 (37.2%) |  |
| Now |  | 37,139,463 (19.3%) | 36,134,283 (19.3%) | 1,005,180 (19.6%) |  |
| BMI | 191,205,305 | 29.12 ± 6.91 | 29.03 ± 6.86 | 32.61 ± 7.87 | <0.001 |
| Waist (cm) | 188,150,185 | 99.09 ± 16.51 | 98.79 ± 16.38 | 110.21 ± 17.41 | <0.001 |
| CVD, n (%) | 192,426,982 | 13,776,816 (7.2%) | 12,511,132 (6.7%) | 1,265,684 (24.7%) | <0.001 |
| Hypertension, n (%) | 192,426,982 | 55,851,108 (29.0%) | 52,645,056 (28.1%) | 3,206,052 (62.5%) | <0.001 |
| Hyperlipidaemia, n (%) | 174,166,347 | 59,000,325 (33.9%) | 56,265,750 (33.2%) | 2,734,576 (55.4%) | <0.001 |
| Diabetes, n (%) | 192,426,982 | 16,810,847 (8.7%) | 15,648,533 (8.4%) | 1,162,314 (22.6%) | <0.001 |
| Nephrolithiasis, n (%) | 192,426,982 | 17,329,578 (9.0%) | 16,557,608 (8.8%) | 771,969 (15.0%) | <0.001 |
| HUA, n (%) | 192,426,982 |  |  |  | <0.001 |
| Non-HUA |  | 158,705,924 (82.5%) | 156,242,184 (NA%) | 2,463,740 (NA%) |  |
| HUA |  | 33,721,057 (17.5%) | 31,052,819 (NA%) | 2,668,239 (NA%) |  |
| Creatinine (μmol/L) | 192,424,374 | 77.26 ± 29.87 | 76.69 ± 26.26 | 97.89 ± 88.64 | <0.001 |
| Serum glucose (mmol/L) | 192,426,982 | 5.48 ± 1.84 | 5.45 ± 1.80 | 6.43 ± 2.86 | <0.001 |
| eGFR | 192,424,374 | 96.24 ± 21.52 | 96.70 ± 21.32 | 79.37 ± 22.03 | <0.001 |
| Cholesterol (mmol/L) | 192,426,982 | 5.04 ± 1.07 | 5.03 ± 1.07 | 5.07 ± 1.18 | 0.6 |
| Triglyceride (mmol/L) | 192,358,986 | 1.74 ± 1.44 | 1.72 ± 1.42 | 2.35 ± 1.90 | <0.001 |
| Direct HDL cholesterol (mmol/L) | 192,401,422 | 1.38 ± 0.42 | 1.38 ± 0.42 | 1.23 ± 0.39 | <0.001 |
| Uric Acid (umol/L) | 192,426,982 | 322.51 ± 83.91 | 320.02 ± 81.89 | 413.28 ± 103.94 | <0.001 |
| Vitamin_C (mg) | 192,426,982 | 81.47 ± 74.24 | 81.81 ± 74.50 | 69.12 ± 62.74 | 0.003 |
| VCQ, n (%) | 192,426,982 |  |  |  | 0.2 |
| Q1 |  | 48,001,038 (24.9%) | 46,523,048 (24.8%) | 1,477,991 (28.8%) |  |
| Q2 |  | 48,269,722 (25.1%) | 46,800,328 (25.0%) | 1,469,394 (28.6%) |  |
| Q3 |  | 48,115,700 (25.0%) | 46,929,892 (25.1%) | 1,185,808 (23.1%) |  |
| Q4 |  | 48,040,521 (25.0%) | 47,041,735 (25.1%) | 998,786 (19.5%) |  |

<sup>1</sup>Mean ± SD or n (%)
<sup>2</sup>Wilcoxon rank-sum test for complex survey samples; chi-squared test with Rao & Scott's second-order correction
Abbreviations: eGFR, glomerular filtration rate; HUA, hyperuricemia

#### 3.1.2 Relationship Between HUA/Gout and VC Intake Level

Table 3 demonstrates the relationship between HUA/gout and VC intake level quartile. Logistic regression showed that Model 1, which did not correct for any variables, had the weakest correlation between VC intake level and the risk of HUA/gout. Model 2 increased the association between VC intake and the risk of HUA/gout; it controlled for age, sex, race, marital status, education, alcohol consumption, and smoking-related demographic variables. Model 3 corrected all covariates. After correction, the correlation between VC intake and the risk of HUA and gout in Model 3 was stable. Additionally, in these three models, VC intake and the risk of HUA in Q4 were generally lower than in Q1–3. After correcting for covariates, the protective effects of VC became increasingly evident from Model 1 to Model 3.

**Table 3.** Association between dietary vitamin C levels (quartiles) and HUA or gout.

| Disease |  |  | Vitamin C |  |  |  | P-value for trend |
| --- | --- | --- | --- | --- | --- | --- | --- |
|  |  |  | Q1 | Q2 | Q3 | Q4 |  |
| HUA | Model 1 | OR (95% CI) | — | 0.80 (0.70–0.91) | 0.82 (0.72–0.94) | 0.68 (0.59–0.78) | <0.001 |
|  |  | P-value | — | 0.001 | 0.005 | <0.001 |  |
|  | Model 2 | OR (95% CI) | — | 0.78 (0.68–0.89) | 0.81 (0.70–0.93) | 0.65 (0.56–0.76) | <0.001 |
|  |  | P-value | — | <0.001 | 0.004 | <0.001 |  |
|  | Model 3 | OR (95% CI) | — | 0.81 (0.68–0.96) | 0.90 (0.76–1.06) | 0.78 (0.65–0.93) | <0.05 |
|  |  | P-value | — | 0.014 | 0.2 | 0.007 |  |
| Gout | Model 1 | OR (95% CI) | — | 0.99 (0.64–1.52) | 0.80 (0.53–1.20) | 0.67 (0.46–0.97) | <0.05 |
|  |  | P-value | — | >0.9 | 0.3 | 0.034 |  |
|  | Model 2 | OR (95% CI) | — | 0.98 (0.63–1.53) | 0.74 (0.47–1.16) | 0.63 (0.42–0.94) | <0.05 |
|  |  | P-value | — | >0.9 | 0.2 | 0.025 |  |
|  | Model 3 | OR (95% CI) | — | 1.03 (0.65–1.63) | 0.83 (0.52–1.34) | 0.77 (0.50–1.20) | >0.05 |
|  |  | P-value | — | 0.9 | 0.4 | 0.3 |  |

Abbreviations: HUA, hyperuricemia

#### 3.1.3 Dose–response Relationship Between HUA/Gout and VC Intake

After completely adjusting for confounding factors and correcting for all covariates, restricted cubic spline analysis was performed for HUA/gout and VC intake, respectively. This showed that VC intake had a non-linear relationship with both HUA (p < 0.0001) and gout (p = 0.001) in participants affected by either (Figure 2). Among these participants, a VC intake of 75–525 mg and 75–225 mg can effectively target HUA and gout, respectively.

**Figure 2.**
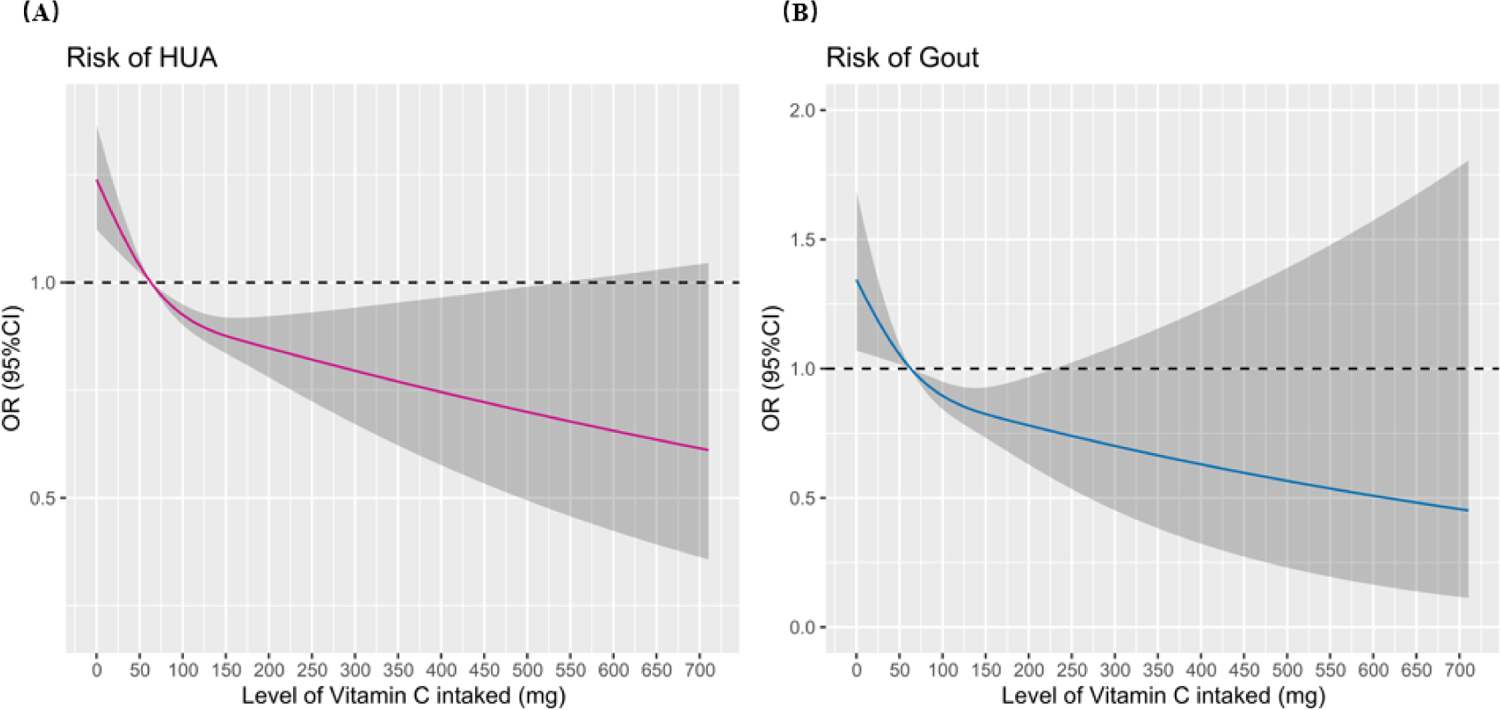
Restricted cubic spline analysis of the association between VC intake and HUA or Gout

### 3.2 Bi-directional MR Study

#### 3.2.1 MR Analysis Using IVs Based on GWAS

SNPs were selected according to the following criteria: SNPs related to confounding factors between VC and HUA were excluded (e.g. those related to haemolysis, diabetes, cholesterol, lymphoma, chemotherapy, alcohol, metabolic syndromes, metabolism, and gout); SNPs related to confounding factors between VC and gout were excluded (e.g. those related to diabetes, cholesterol, alcohol, metabolic syndromes, metabolism, uric acid levels, HUA, and hypertension); SNPs related to the results; and SNPs that were not present in the results. For MR analysis, we selected all SNPs with critical values of p < 1 × 10^-5^ related to HUA/gout–VC absorbates; the remaining selection (p < 1 × 10^-6^) was used as an IV to increase the number of available SNPs and to facilitate subsequent sensitivity analyses.

We assessed the causal relationship between VC and HUA/gout based on different IVs (Table 4). The MR results show a causal relationship between VC absorbates and HUA; the MR IVW results show a negative correlation between VC absorbates and HUA (OR = 0.985, 95% CI = 0.973–0.997, p = 0.015). The results of all three other methods, MR-Egger, weighted median, and weighted mode, were statistically significant (p < 0.05), consistent with the IVW results. Figure 3A shows a scatterplot of the correlation between SNPs and HUA, as well as between SNPs and VC absorbates. Figure 3B shows a forest plot for each SNP in VC absorbate–HUA estimation, suggesting the magnitude of the MR effects of VC on HUA. As shown in Table 4, no statistically significant relationship was observed between supplemental VC or pharmacological VC and HUA in this MR analysis.

**Figure 3.**
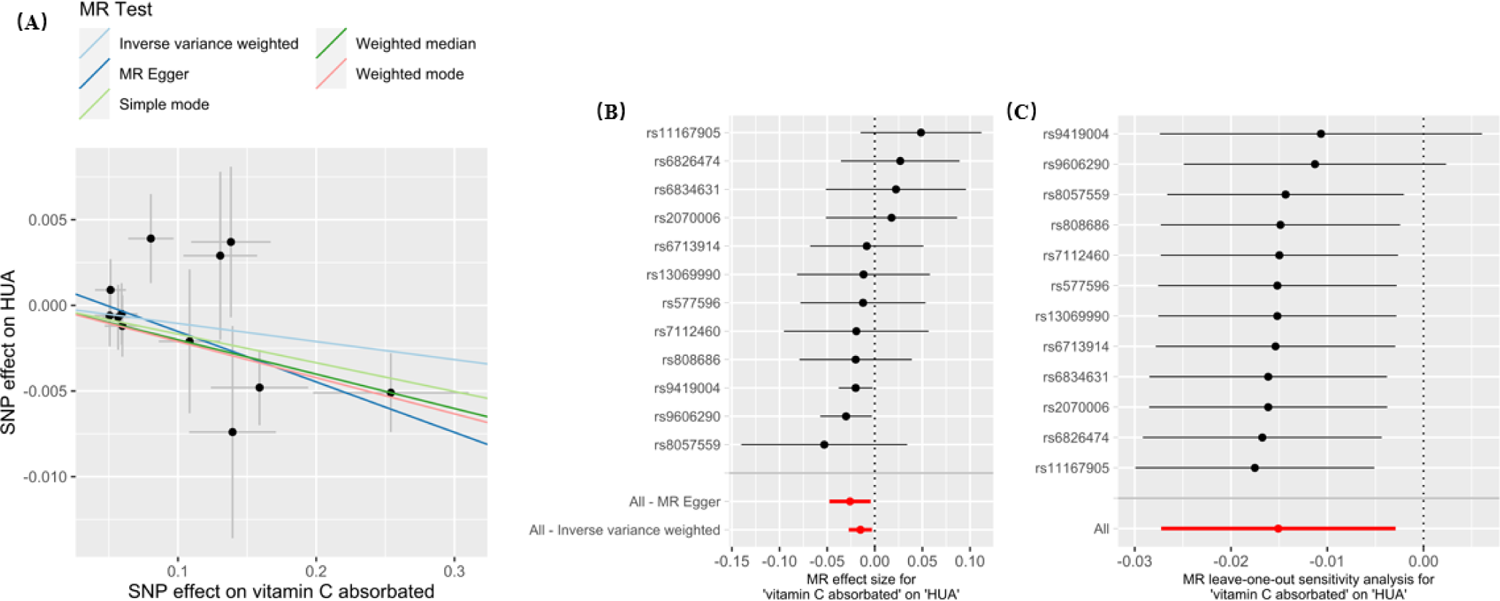
MR Effect of vitamin C absorbated on HUA, Note. (A) Scatter plot of SNP-HUA association versus SNP-vitamin C absorbated association; (B) Forest plots of vitamin C absorbated-HUA estimates for each SNP; (C) Leave-one-out sensitivity analysis.

**Table 4.**
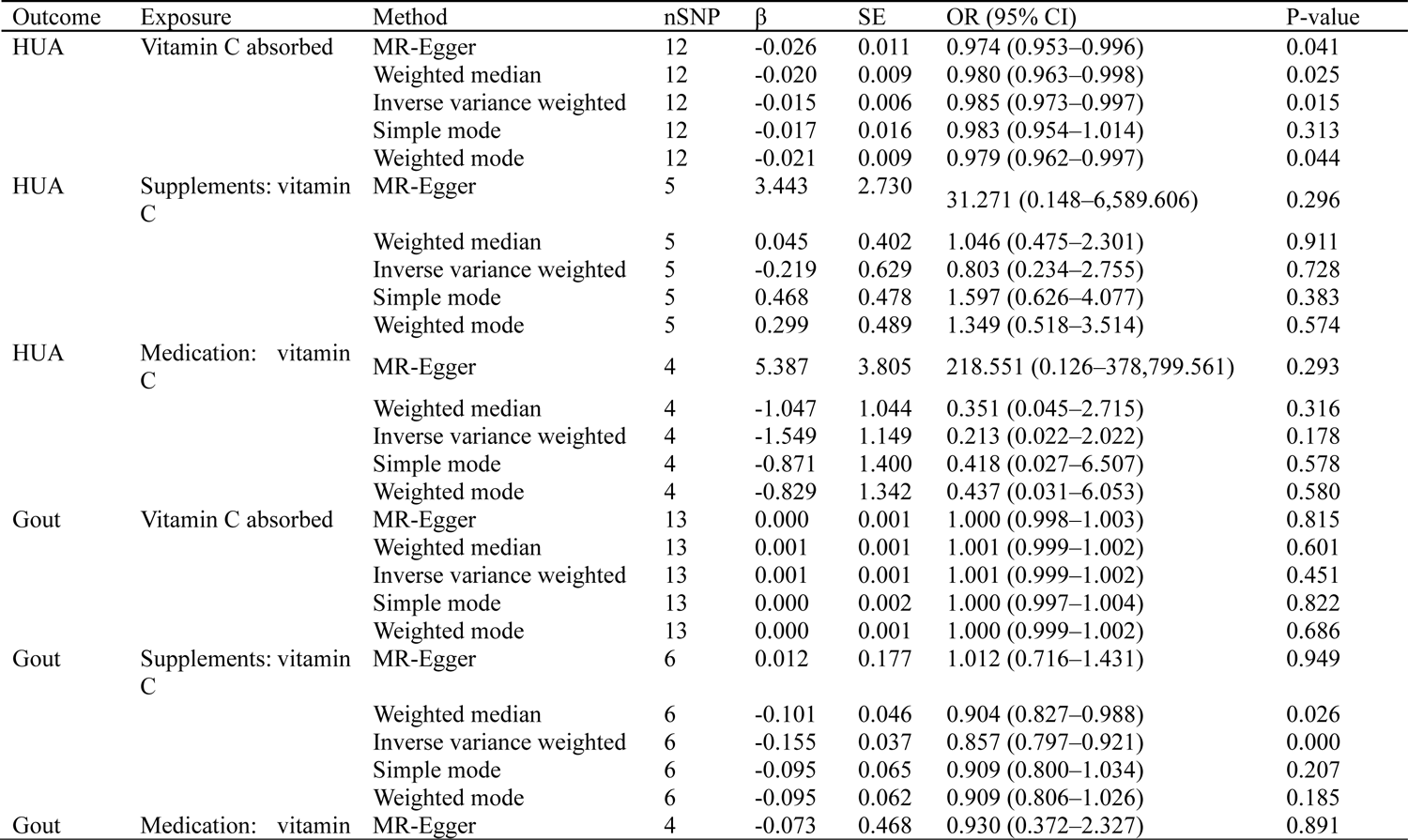
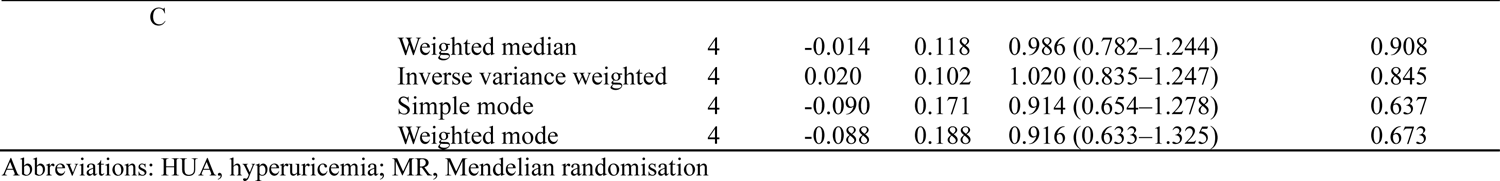
MR results for the causal effect of vitamin C on HUA and gout.

In addition, the MR IVW analysis results show a causal relationship between supplemental VC and gout (OR = 0.857, 95% CI = 0.797–0.921, p < 0.001), supported by the weighted median method (p < 0.05); correlation scatter plots and forest plots indicate the magnitude of the MR effect of VC on gout, as shown in Figure 4A and B. However, there was no statistically significant relationship between supplemental VC absorbates or pharmacological VC and gout in the MR analysis, as shown in Table 4.

**Figure 4.**
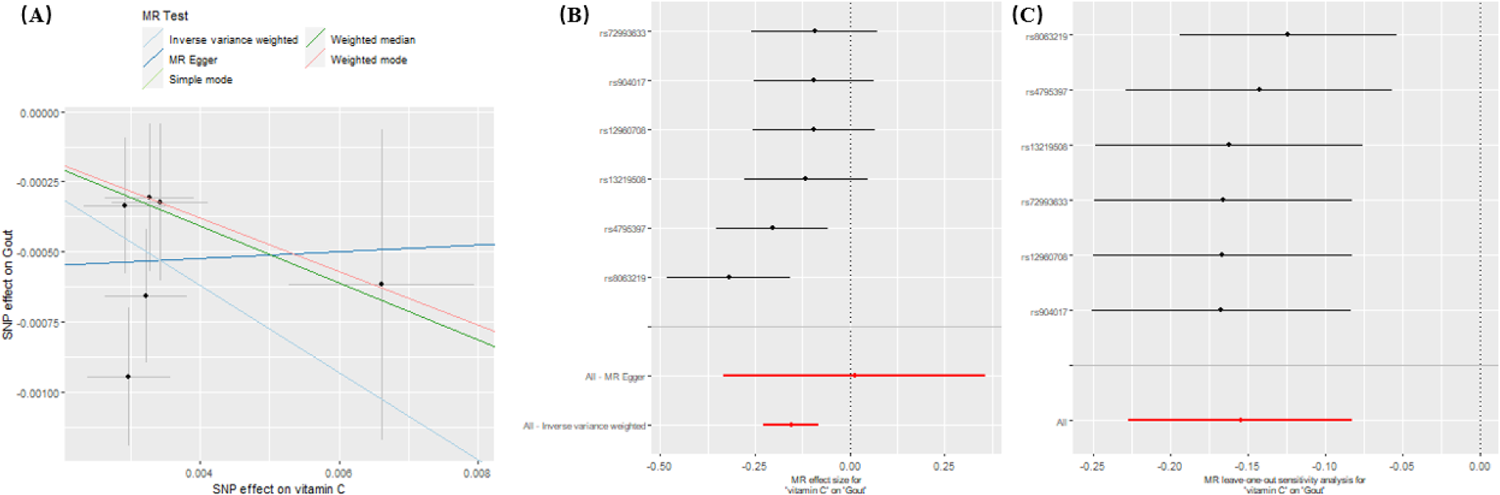
MR Effect of supplements: vitamin C on Gout, Note. (A) Scatter plot of SNP-Gout association versus SNP-vitamin C absorbated association; (B) Forest plots of vitamin C absorbated-Gout estimates for each SNP; (C) Leave-one-out sensitivity analysis.

#### 3.2.2 MR Sensitivity Analysis

We estimated the stability of the MR results using sensitivity analysis, as shown in Table 5. In the analysis of VC absorbates versus HUA, Cochran’s Q test showed that there was slight heterogeneity in the MR-Egger analysis (p = 0.049). The MR-Egger intercept test indicated that the MR analyses were not affected by horizontal pleiotropy (p > 0.05). Leave-one-out sensitivity analysis confirmed the robustness of the MR results, and the absence of major SNPs (Figure 3C) greatly affected the results when eliminated. In addition, for the analysis of supplemental VC versus gout, Cochran’s Q test showed no heterogeneity in the MR-Egger analysis (p > 0.05), and the results of the MR-Egger intercept test indicated that the MR analysis was not affected by horizontal pleiotropy (p > 0.05). Leave-one-out sensitivity analysis showed no major SNPs that would greatly affect the results when eliminated, confirming the robustness of the MR results, as shown in Figure 4C.

**Table 5.**
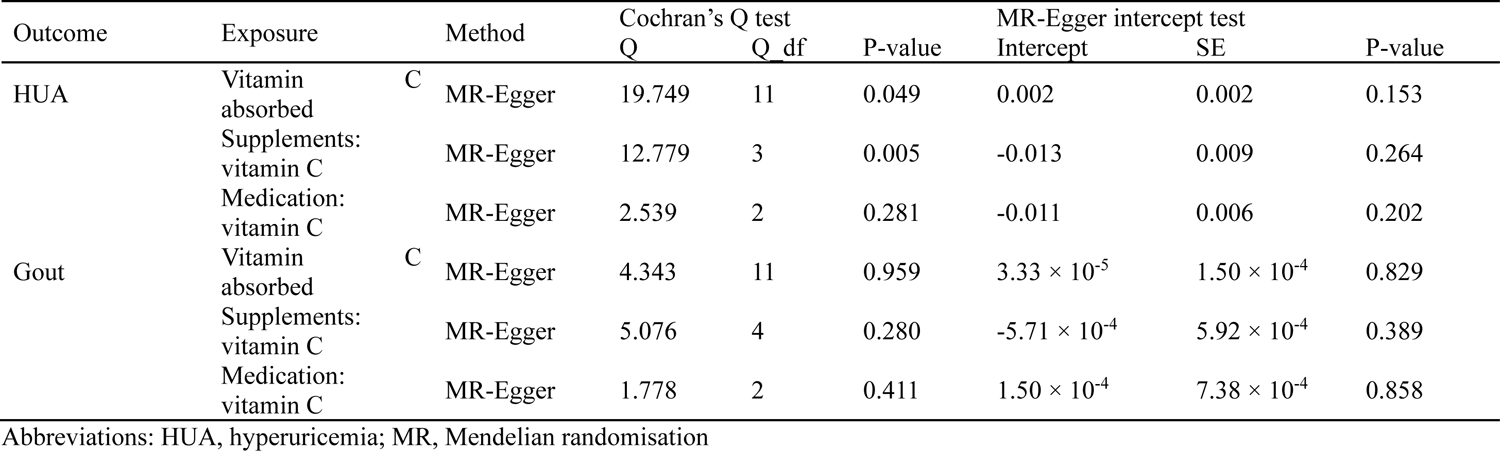
Sensitivity analysis between vitamin C and HUA and gout.

#### 3.2.3 Reverse MR Analysis of the Causal Relationship Between VC and HUA/Gout

We performed reverse MR analysis to estimate whether VC would affect HUA/gout using SNPs with critical values of p < 5 × 10^-8^ as IVs. The reverse MR results are shown in Table 6. The results show that there is no reverse causal relationship between absorbed VC and HUA (p > 0.05 for all five MR methods). Additionally, there was no reverse causal relationship between supplemental VC and gout (p > 0.05 for all five MR methods).

**Table 6.**
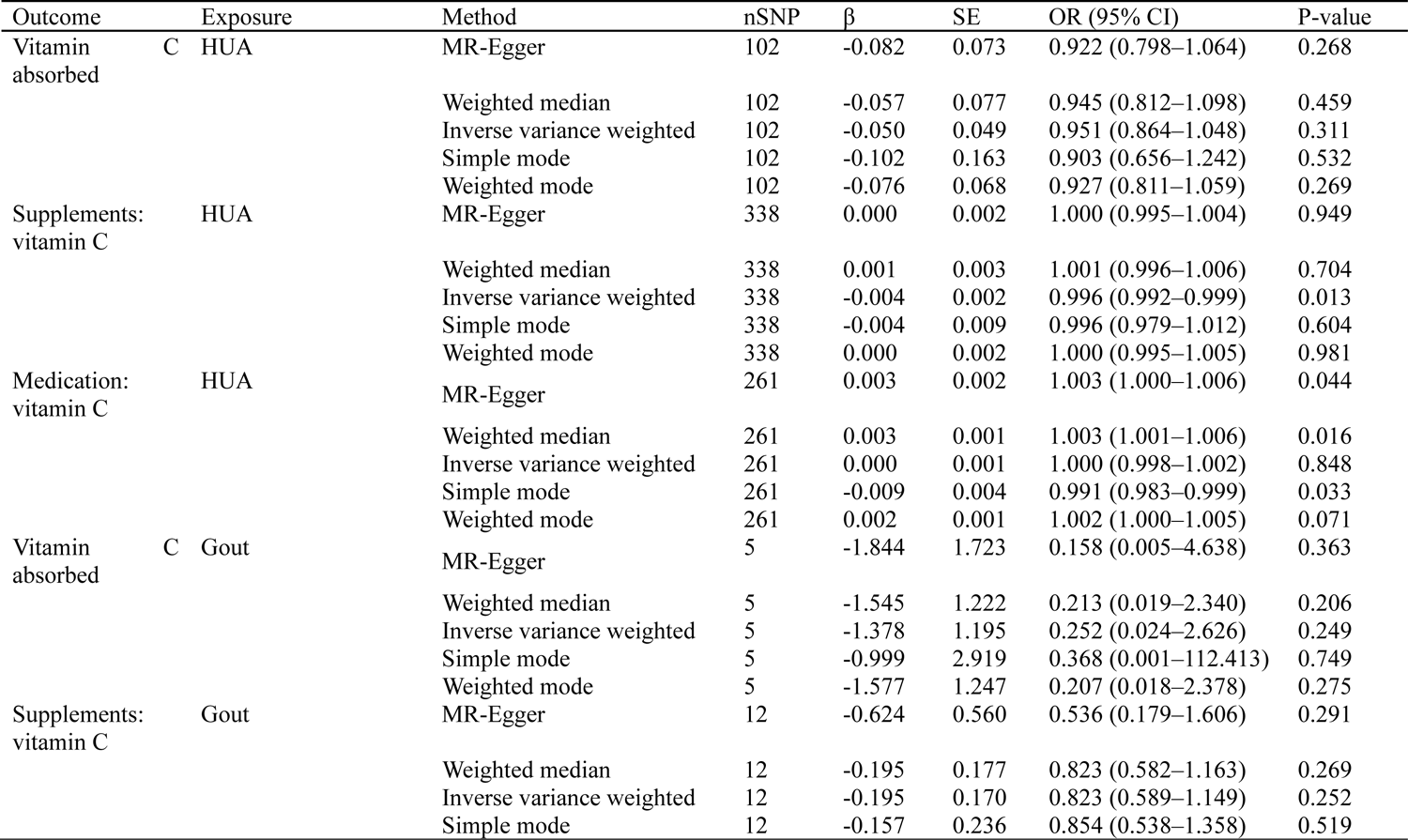
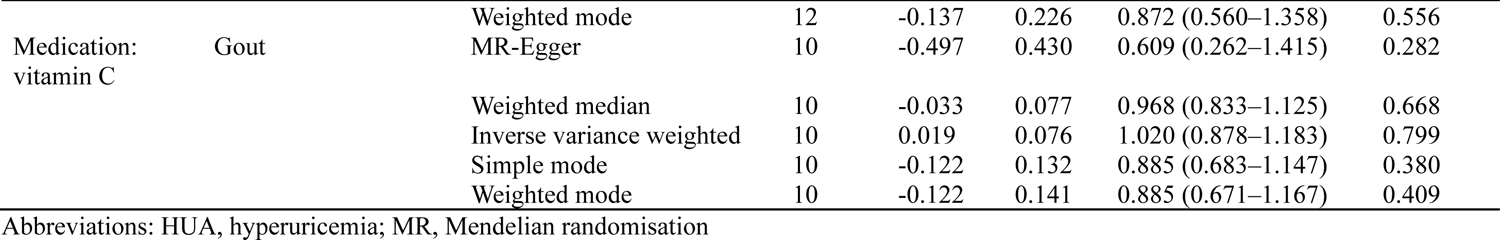
Results for the causal effect of vitamin C on HUA and gout.

Four MR methods exhibited no reverse causal relationship between supplemental VC and HUA (p > 0.05), whereas two methods showed no reverse causal relationship between pharmacological VC and HUA (p > 0.05). Each MR method found no reverse causal relationship between any form of VC and gout (p > 0.05).

#### 3.2.4 Reverse MR Sensitivity Analysis

Cochran’s Q test showed no heterogeneity in the reverse MR analysis of absorbed VC and HUA (p > 0.05), whereas there may be slight heterogeneity in the reverse MR analysis of supplemental VC and gout (p = 0.035). In addition, the MR-Egger intercept test showed that the inverse MR analyses of absorbed VC versus HUA and supplemental VC versus gout were not affected by pleiotropy (p > 0.05) (Table 7). Finally, leave-one-out sensitivity analysis confirmed the robustness of the reverse MR results (Supplementary Figures 1 and 2).

**Table 7.**
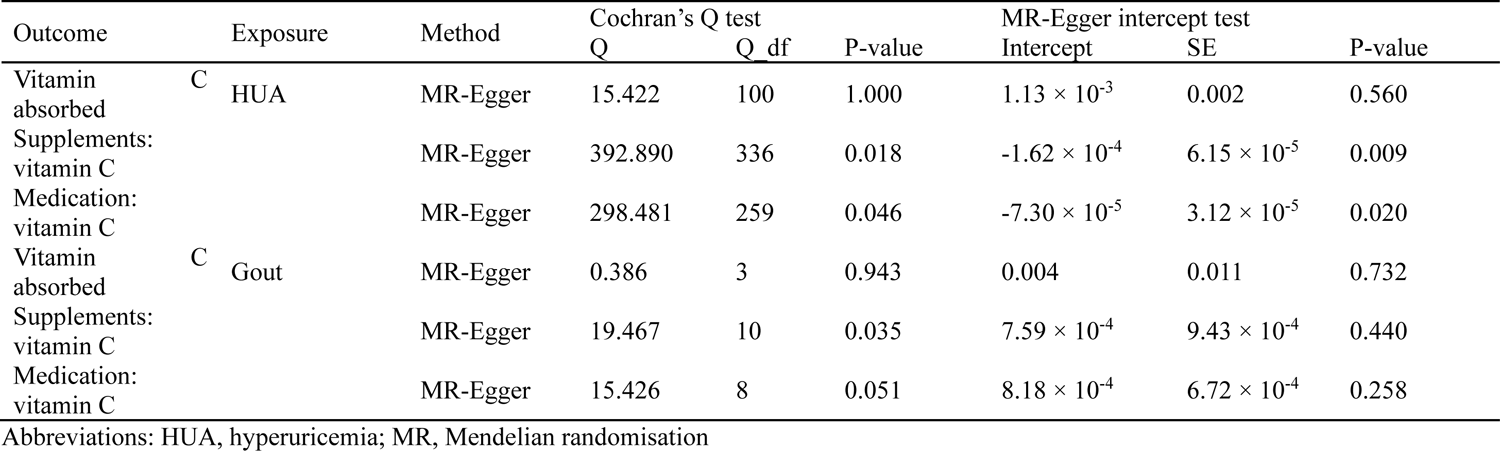
Sensitivity analysis between vitamin C and HUA and gout.

## 4 Discussion

There is evidence from prospective epidemiological studies that dietary factors are clinically associated risk factors for HUA and gout, including evidence for the protective effects of VC (15). Some researchers considered that the urate-lowering effect of VC seemed to be more important than other functions. They further explored the potential mechanism of its urate-lowering properties and found that VC may reduce urate-induced inflammation by inhibiting the activation of the NLRP3 inflammasome (16). Additionally, in vivo studies showed that the VC transporter SVCT1 acts as a urate importer and that SVCT1 damage can lower serum uric acid levels (17). However, studies that confirm the relationship between dietary VC intake, serum uric acid concentration, and gout are lacking.

In this study, we used data from the 2007–2018 NHANES cross-sectional survey and performed bi-directional MR analysis to accurately investigate the relationship between gout/HUA risks and dietary VC intake levels. The results show that gout/HUA and VC intake are negatively correlated in the cross-sectional survey. Restricted cubic spline analysis showed that daily dietary intakes of 75–525 mg and 75–225 mg VC have protective effects on patients with HUA and gout, respectively. In the MR analysis, the causal association between absorbed VC and HUA, as well as supplemental VC and gout was consistent with the cross-sectional study.

Consistent with the present study, the dietary and nutritional factors associated with HUA in the Seventh Korean NHANES showed that dietary fibre intake and VC intake were negatively correlated with HUA (18). Another case-control study on the association between diet/obesity and gout in Taiwan showed that low VC intake increased the risk of gout and that increasing food sources rich in VC and dietary fibre can prevent gout (19). A meta-analysis of randomised controlled trials showed that a median supplementary VC dose of 8 mg/day for at least one week significantly decreased HUA (20). In contrast, current clinical dietary interventions that can reduce the frequency of gout flares (e.g. VC supplementation) can reduce serum uric acid levels and be used as adjuncts for urate-lowering treatment, a non-pharmacological approach to treat gout and HUA (21).

However, further research shows that in patients with gout, continuous VC treatment for eight weeks did not obviously lower uric acid levels, whether this constituted VC alone (500 mg/d) or VC combined with allopurinol. The results of this study may be biased due to a variety of factors such as dose, participants, and course of treatment (22). Overall, neither clinical observational studies nor randomised controlled trial meta-analyses can control for confounding factors and bias. Furthermore, most clinical studies are small-scale randomised controlled trials, which cannot control the accuracy of the results. Additionally, interventional studies of dietary VC intake levels and HUA/gout are still lacking, necessitating further research.

MR analysis can appropriately supplement deficiencies when using a large-sample GWAS database without fixed periods and can predict the permanent effects of VC intake. As the current database did not have data related to dietary VC intake, in the present study, it was important to use MR to explore the causal relationship between the risk of HUA/gout and supplemental VC, pharmacological VC, and ascorbic acid, respectively. In the MR results, the genetic susceptibility of absorbed VC with HUA and supplemental VC with gout is not coincidental; forward analysis and reverse MR testing also supported these negative correlations. The potential mechanisms of action underlying these effects of VC can include the following.

First, a gene mutation during the evolution of humans and guinea pigs means that endogenous VC cannot be produced. In addition, VC maintains homeostasis through anti-oxidation and cofactor functions. Blood lacks VC and easily undergoes oxidative stress and peroxide damage, but humans and guinea pigs need endogenous supplementation (23, 24). Second, the most important physiological process in which VC is involved is anti-oxidation. According to research findings, another genetic mutation has been discovered in humans, leading to the elimination of the active uricase gene. This gene can counteract VC deficiency by increasing serum uric acid levels. High intracellular uric acid levels are prone to inducing oxidative stress in the body, and VC reduces the production of mitochondrial Reactive Oxygen Species, increases the level of antioxidant enzymes, and enhances the ability of the body to dissolve uric acid and promote uric acid efflux (25).

However, the VC transporter protein modifies the function and activity of URAT1 in proximal renal tubular cells, thus increasing urate excretion in the body. As the body may obtain more fructose than other nutrients during VC intake, the role of VC; however, it will increase the level of uric acid, gout risk, and thus has obvious limitations. A side-by-side study evaluating dietary factors and biochemical indices of patients with gout and asymptomatic HUA confirmed that serum uric acid levels are influenced by dietary factors, and that VC intake in the healthy group was lower than in the disease group. This suggests that dietary intake of energy, protein, and fructose also affects gout and HUA (26). The lack of a significant correlation of supplements: vitamin C and medication: vitamin C with the risk of HUA, and vitamin C absorbated and medication: vitamin C with the risk of gout in the MR study could be attributed to these factors.

There are several distinct advantages to the present study. First, not only did this study utilise the NHANES database spanning the longest period, 2007–2018, for data completeness, but it also allowed for an investigation of the association between dietary VC intake and gout/HUA at the epidemiological level. Second, we introduced bi-directional MR analysis, which can address unavoidable influencing factors, such as reverse causality, measurement error, and residual confounders, which exist in ordinary epidemiological studies as well as NHANES data analysis; this adds practical significance to this study. One of the key aspects of this study is its inclusive examination of both gout and HUA, deviating from the conventional approach of focusing solely on a single factor, thus facilitating a more comprehensive investigation of the complexity of this type of immune disease. It facilitated the comparison between each disease to provide more informed advice based on patients in the clinic, as well as experimental direction for researchers to conduct further in vivo and in vitro studies. However, certain limitations exist, including the reliance solely on screening and analysis from the intrinsic database and the absence of extensive, prolonged multi-centre clinical research involving a larger sample size. Further research is required to validate the findings through long-term observational and interventional studies.

In conclusion, our study supports a causal relationship between dietary VC intake and the risk of HUA, with cross-sectional studies as well as a portion of our MR analyses suggesting a significant negative association. However, despite observational studies suggesting a negative association, other MR analysis did not yield significant results, suggesting that the results might still be influenced by uncontrollable confounding factors. Therefore, based on our findings, we hypothesise that daily dietary VC intake doses of 75–525 mg and 75–225 mg may potentially decrease the risk of HUA and gout, respectively; however, further studies with larger sample sizes are necessary to confirm these findings.

## Abbreviations

GWAS: genome-wide association study

HUA: hyperuricemia

IV: instrumental variable

IVW: inverse variance weighting

MR: Mendelian randomisation

NHANES: National Health and Nutrition Examination Survey

SNP: single nucleotide polymorphism

VC: vitamin C

## 7 Author Contributions

Z-NP and X-QW conceptualized the study, analyzed and interpreted the data, and revised and edited the manuscript. QD, W-TY, W-QZ, Y-BP, NL, J-YP and L-LG drafted the manuscript. NL, J-YP QD, W-TY and W-QZ provided advice for the project. NL, J-YP and L-LG provided administrative support. All authors read and approved the final manuscript.

## 9 Acknowledgments

The authors wish to acknowledge the authors and participants of the involved GWAS for providing summary statistics data.

## 11 Data Availability Statement

The raw data supporting the conclusions of this article will be made available by the authors, without undue reservation.

All data produced in the present study are available upon reasonable request to the authors. All data produced in the present work are contained in the manuscript. All data produced are available online at.

## Footnotes

1 https://shiny.cnsgenomics.com/mRnd/

