## Supplement table 1and Figure 1∽2 for "Analysis of dietary vitamin C intake levels and the risk of hyperuricemia and gout based on cross-sectional studies and bi-directional Mendelian randomisation"

Supplement table1 Characteristics of data in this study

| Outcome/Exposure | Trait | Sample size | Population | Number of SNPs | Dataset | Data source |
|---|---|---|---|---|---|---|
| vitamin C absorbated | Ascorbate (Vitamin C) | 2,085 | European | 2,545,101 | met-a-348 | GWAS summary data https://gwas.mrcieu.ac.uk/ （PMID: 24816252） |
| supplements: vitamin C | Vitamin and mineral supplements: Vitamin C | 460,351 | European | 9,851,867 | ukb-b-15175 | GWAS summary data https://gwas.mrcieu.ac.uk/ Note: 6155#3: Output from GWAS pipeline using Phesant derived variables from UKBiobank |
| medication: vitamin C | Treatment/medication code: vitamin c product | 462,933 | European | 9,851,867 | ukb-b-488 | GWAS summary data https://gwas.mrcieu.ac.uk/ Note: 20003# 1140909726: Output from GWAS pipeline using Phesant derived variables from UKBiobank |
| HUA | Serum uric acid levels | 343,836 | European | 19,041,286 | ebi-a-GCST90018977 | GWAS summary data https://gwas.mrcieu.ac.uk/ （PMID: 34594039） |
| Gout | Gout | 484,598 | NA | 9,587,836 | ebi-a-GCST90038687 | GWAS summary data https://gwas.mrcieu.ac.uk/ （PMID: 33959723） |

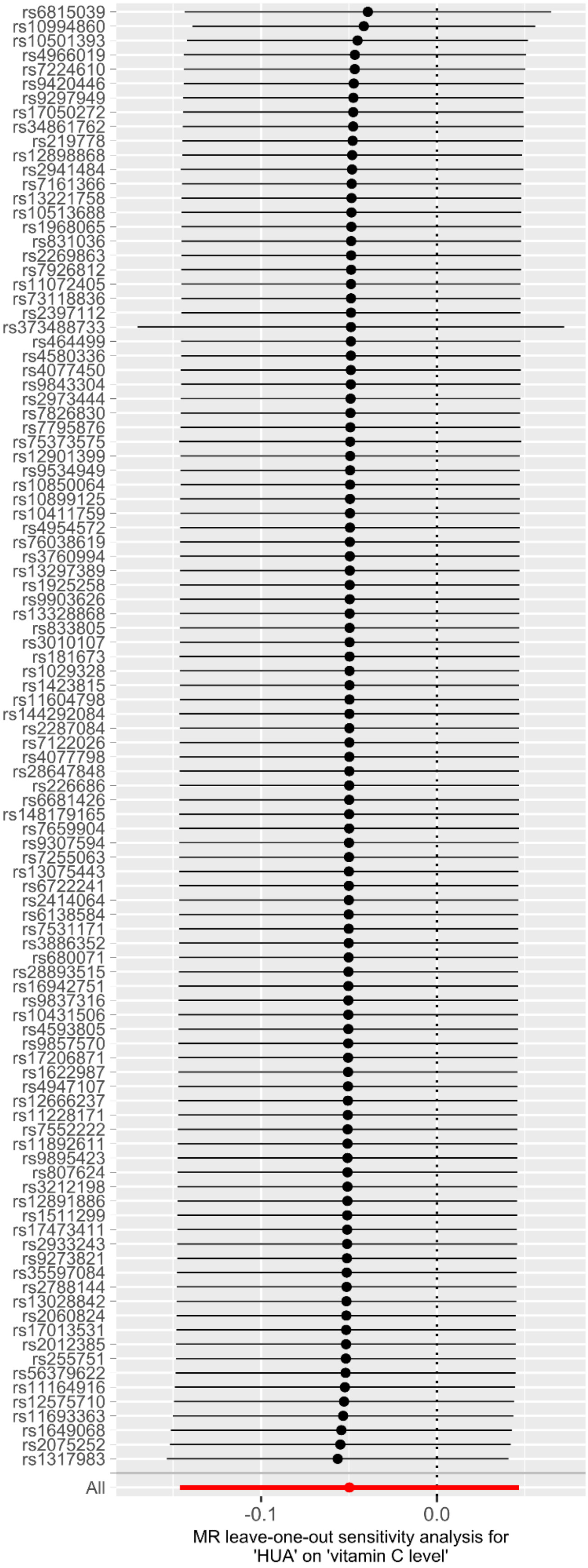


Supplement Figure 1. Sensitivity analysis of HUA and vitamin C absorbated

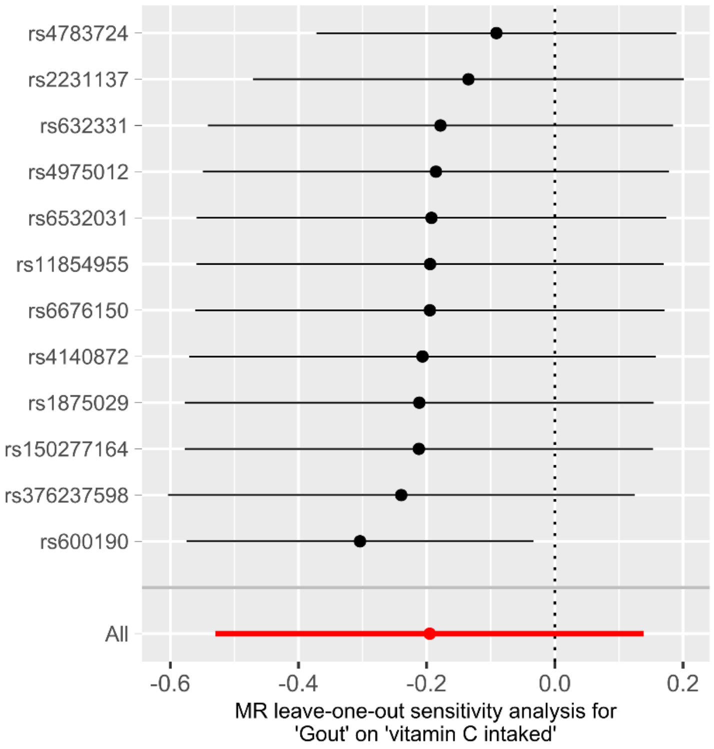


Supplement Figure 2. Sensitivity analysis of Gout and supplements: vitamin C
